# Quantitative T1 mapping indicates elevated white matter myelin in children with RASopathies

**DOI:** 10.1101/2025.02.25.25322881

**Authors:** Julia R. Plank, Elveda Gozdas, Jennifer Bruno, Chloe A. McGhee, Hua Wu, Mira M. Raman, Manish Saggar, Tamar Green

**Affiliations:** Division of Interdisciplinary Brain Sciences, Department of Psychiatry and Behavioral Sciences, 1520 Page Mill Road, Palo Alto, CA 94304, USA; Center for Cognitive and Neurobiological Imaging, Stanford University, Stanford, CA 94305, USA

**Keywords:** quantitative T1 mapping, myelin, neurofibromatosis type 1, Noonan syndrome, RASopathies, magnetic resonance imaging

## Abstract

**Background:** Evidence suggests pathological roles of myelination in neurodevelopmental disorders, but our understanding is limited. We investigated quantitative T1 mapping (QT1) as a clinically feasible tool for measuring myelination in children with neurodevelopmental disorders of the RAS-MAPK signaling pathway (RASopathies).

**Methods:** We collected QT1, diffusion-weighted, and structural MRI scans from 72 children (49 RASopathies, 23 typical developing (TD)). QT1 measures of myelin content included the macromolecular tissue volume (MTV) in white matter and R1 (1/T1 relaxation) of the cortex. For white matter, we assessed between-groups differences across 39 tracts. For cortical R1, we used principal components analysis to reduce dimensionality and capture myelination patterns across 360 regions. A multivariate ANOVA assessed differences across principal components. Finally, a support vector machine (SVM) identified the most discriminative features between TD and RASopathies.

**Results:** Thirty-four of 39 tracts were higher in MTV in RASopathies relative to TD (*p_FDR_*<.05), indicating widespread elevation in myelination. Our MANOVA revealed a group effect on cortical R1 (*p*=.002, *η^2^*=.028), suggesting cortical myelination differences between-groups. SVM yielded an accuracy of 87% and identified cognitive and cortical R1 features as the most discriminant between-groups.

**Conclusions:** We found widespread elevated myelin in white matter tracts and region-dependent patterns of cortical myelination in children with RASopathies. QT1 enabled us to leverage preclinical models showing oligodendrocyte dysfunction to uncover the myelination pattern *in vivo* in the developing human brain. Using QT1, myelin represents a promising treatment target that can be identified and monitored in neurodevelopmental disorders, offering significant potential for advancing current therapeutic strategies.

## Introduction

Neurodevelopmental disorders affect an estimated 15% of children and adolescents worldwide (1); however, effective treatments are limited due to a poor understanding of the underlying pathology. Many of these conditions, such as attention-deficit hyperactivity disorder (ADHD) and autism spectrum disorder (ASD), are associated with aberrations in white matter myelin (2). Consequently, recent work emphasizes the importance of understanding myelin biology (3), particularly as myelin is linked to cognitive functioning (4) and, therefore, may serve as a therapeutic target in future clinical trials. However, interpreting myelin abnormalities in neurodevelopmental disorders *in vivo* presents significant challenges due to the heterogeneity of these conditions and the lack of specific neuroimaging techniques capable of accurately visualizing myelin integrity and composition.

The heterogeneity associated with neurodevelopmental disorders can be mitigated through the adoption of a genetics-first approach. Thus, this study focused on disorders caused by single germline mutations in the RAS-extracellular signal-regulated mitogen-activated protein kinase (RAS-MAPK) signaling pathway. By studying disorders with a clear genetic basis, we can also leverage preclinical models to inform our results. Animal models previously showed that oligodendrocytes, the myelin-producing glial cells, are affected in common RAS-MAPK disorders (collectively termed ‘RASopathies’) such as Noonan syndrome and neurofibromatosis type 1. Causal mutations of these RASopathies upregulate the RAS-MAPK pathway and lead to downstream cellular effects including proliferation of oligodendrocyte precursor cells and fewer myelinated axons (5). Gaining insight into the brain pathology of RASopathies may also shed light on further neurodevelopmental disorders. Notably, ADHD and ASD are frequent comorbidities in this population (6,7) and emerging genetic evidence suggests the RAS-MAPK pathway may be involved in a variety of currently ‘idiopathic’ neurodevelopmental disorders (8). By utilizing a genetics-first approach to investigate syndromes with well-established genetic underpinnings, we can generate valuable models for improving our understanding of myelin changes across various neurodevelopmental disorders.

Previous *in vivo* neuroimaging studies of neurodevelopmental disorders have primarily relied on diffusion tensor imaging (DTI) to make inferences about white matter abnormalities. While DTI is sensitive to changes in white matter, these findings are generally non-specific (9). For example, decreased fractional anisotropy (FA) may be caused by reduced neurite density, demyelination, increased orientation dispersion, or other factors (10). In contrast, quantitative T1 (QT1) mapping is an MRI technique that provides specific measures of brain tissue composition, particularly myelin content (11). Furthermore, QT1 is robust to differences in scanner hardware and boasts short acquisition times averaging around 3 minutes, making it particularly suitable for pediatric populations. QT1 utilizes the MR signal from protons to reliably assess the macromolecular content of tissue, i.e., cell membranes, proteins, and lipids contained within myelin sheaths. Previous work showed that the R1 (1/T1 relaxation) and macromolecular tissue volume (MTV) measurements derived from QT1 are accurate indicators of myelin content (11,12). By utilizing cortical R1 and white matter tract MTV, we can acquire a comprehensive estimation of myelin content changes throughout the brain in neurodevelopmental disorders.

In this study, we investigated the application of QT1 mapping for measuring myelin content in children with RASopathies compared to their typical developing (TD) peers. To our knowledge, this represents the first study to apply QT1 mapping to children with RASopathies. We hypothesized that alterations to R1 and MTV would be found in the RASopathies cohort compared to TD, suggesting significant differences in myelination. We expected to find shorter R1 and smaller MTV values, given that preclinical work has suggested hypomyelination in mouse models of Noonan syndrome (5). We utilized univariate and multivariate analyses to discern group differences but also employed SVM to pinpoint the most discriminative features separating TD children from those with RASopathies. Our study builds on previous DTI studies of RASopathies (13–17) by using QT1 mapping as a more specific methodology. The potential implications of this investigation include a deeper understanding of the pathology associated with neurodevelopmental disorders, the identification of specific treatment targets, and the translation of QT1 mapping to clinical settings.

## Methods and Materials

### Participants

Participants with RASopathies were recruited for this prospective study from January 2023 through November 2024 across the United States and Canada. TD participants were recruited from the San Francisco Bay area. Eligible participants included 15 children with NF1, 46 with NS, and 27 TD. Full-scale, performance, and verbal IQs were acquired using the Wechsler Abbreviated Scale of Intelligence 2^nd^ Edition (18). Additional cognitive measures included seven tests from the NIH Toolbox Cognition Battery (http://www.nihtoolbox.org/) encompassing measures of attention, memory, and language abilities (19). Further details on these tests and the inclusion and exclusion criteria are available in the **Supplement.** Legal guardians provided written informed consent and participants aged over 7 years provided complementary written assent. The Stanford University School of Medicine Institutional Review Board approved all procedures in this work involving human subjects. All procedures comply with the ethical standards of the national and institutional committees on human experimentation and with the Helsinki Declaration of 1975, as revised in 2008.

### Imaging Protocol

MRI data were acquired using a standard 48-channel head coil on a GE Premier 3.0 Tesla whole-body system (GE Healthcare, Milwaukee, WI). Structural data were collected using a whole-brain high-resolution T1-weighted magnetization-prepared rapid gradient-echo (MPRAGE) sequence. QT1 data were collected using slice-shuffling of an inversion-recovery EPI sequence with multiple inversion times (TI) as previously described (20). We acquired 20 TIs with the first TI = 50 ms and TI interval = 150 ms and a second inversion-recovery EPI with reverse-phase encoding direction. Other scan parameters included: flip angle = 60°, repetition time (TR) = 3.5 s; field-of-view (FOV) = 22 cm; matrix size = 110 x 110; slice thickness = 2.5 mm; number of slices = 57. The sequence generates two QT1 NIFTI files, one with reverse phase encoding for EPI distortion correction. Both NIFTI files were distortion-corrected using *topup* in FSL and were subsequently processed using open-source Python code (https://github.com/cni/t1fit/blob/master/t1fit_unwarp.py) to produce the QT1 maps for further analysis. Diffusion MRI data were collected with *b*=500s/mm^2^ (6 directions), *b*=1000s/mm^2^ (15 directions), *b*=2000s/mm^2^ (15 directions) and *b*=3000s/mm^2^ (60 directions). Preprocessing of diffusion MRI data was completed by author MR using FSL 6.0.5. (FMRIB Analysis Group, Oxford, UK). *Topup* and *eddy* tools were used for susceptibility-induced distortion correction and correction of eddy currents-induced distortions and subject movements (21,22).

### Image Analysis

T1-weighted images were used to reconstruct cortical surfaces for each subject in FreeSurfer (version 5.3, http://surfer.nmr.mgh.harvard.edu). The steps included skull stripping and grey and white matter segmentation followed by reconstruction and inflation of the cortical surface. Manual editing of the segmentation was performed (authors MR and CM) when required. The Human Connectome Project multi-modal parcellation (HCPMMP) (23) was used to delineate 180 cortical brain regions per hemisphere, as described previously (24). Briefly, the HCP annotation files were converted to the standard FreeSurfer cortical surface and the subsequent parcellation was transformed to each participant’s cortical surface. Volumetric masks were generated for each cortical surface, and these were linearly transformed into the native space of each participant’s diffusion-weighted images. Transformations were visually checked for potential artifacts or misalignments using ITK-SNAP. R1 values were extracted from each transformed brain region (360 per subject).

For tract-based analysis, the QT1 maps for each subject were first co-registered to the diffusion-weighted data using the ANTS software package, i.e., each QT1 map was warped to the non-diffusion-weighted b0 image. TRActs Constrained by UnderLying Anatomy (TRACULA) within FreeSurfer was used to reconstruct white matter tracts (25). TRACULA combines the distortion-corrected diffusion MRI data with T1-weighted structural images to reconstruct 42 white matter tracts for each subject using probabilistic tractography. 3D reconstructions of the tracts were visually examined (authors JP, MR) and any tracts that failed or only partially reconstructed were rerun using the *reinit* function. Tracts that failed to reconstruct or did so partially following *reinit* were excluded from analysis. Finally, the average MTV was extracted from each tract using the previously validated equation: 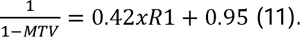

### Statistical Analysis

For tract-based analysis, we used an analysis of covariance to assess between-group differences (RASopathies, TD) in the average tract MTV for each of the 39 white matter tracts, including age and sex as covariates. We used the false discovery rate (FDR) to adjust for multiple comparisons.

For cortical R1 analysis, we performed a non-parametric multivariate ANOVA (MANOVA) to assess for an effect of group (RASopathies, TD) on cortical R1 simultaneously across the 360 regions. Prior to the MANOVA, we regressed out the effects of age on the data. We then performed a PCA on the residuals to reduce the dimensionality. We checked for outlier subjects in the principal components using the Mahalanobis distance for multivariate data. The Mahalanobis distance of each observation was compared to a critical value from the Chi-square distribution. Outliers were identified as those where *p*<.001. The principal components were entered into the MANOVA generated using the *nonpartest* function (10,000 permutations) within the *npmv* package (26). We chose this non-parametric approach as the model is more robust against potential violations of assumptions than the classic parametric MANOVA.

Finally, we investigated the effect of RASopathies on a combination of brain and cognitive features using a support vector machine (SVM) which extracted the most discriminative features between RASopathies and TD. SVM is a machine learning tool that uses a multivariate approach ideal for handling data with a large number of features. SVM classifies the data by finding a hyperplane that maximizes the distance between each class. We used the *fitcsvm* function for binary classification in MATLAB R2024A to fit the SVM for TD versus RASopathies. We entered 406 features including tract MTV, cortical R1, and NIH toolbox subscales, to fit the SVM. Given the small sample size (total n=65 included in SVM analysis), we used a leave-one-out cross-validation to fit the models, i.e., all subjects except one were used to train the model, and then the remaining one subject was used to test the model across 65 iterations. We evaluated the model using receiver operating characteristic (ROC) metrics of accuracy, sensitivity, and specificity. Following the cross-validation, one classifier was produced from which we extracted the ten features with the largest weightings. These features represent the variables with the greatest discriminative power between the classes. Finally, we used permutation-testing (n=1000 permutations) to evaluate our SVM model in comparison to a null model where the subjects were randomly shuffled prior to the leave-one-out cross-validation. Aside from SVM, which was conducted in MATLAB, all other statistical analyses and visualizations were conducted in R 4.4.1 (R Core Team, 2024).

## Results

### Participant Characteristics

A total of 72 subjects (M_age_=10.7, SD_age_=3.47; 36 male) were included in the analysis (**Table 1**). Eleven subjects were excluded due to excess motion (TD=2, RASopathies=9). **Figure 1** shows the flow of participants through the study and reasons for exclusion. There were 49 subjects in the RASopathies group (M_age_=11.0, SD_age_=3.48; 25 male) and 23 subjects in the TD group (M_age_=10.2, SD_age_=3.48; 11 male). The subjects with RASopathies included children with Noonan syndrome (n=34, *PTPN11* (n=20), *SOS-1* (n=8), *NRAS* (n=2), *RAF-1* (n=2), *SHOC-2* (n=1), *LZTR-1* (n=1) and children with neurofibromatosis type 1 (n=15, *NF1* mutation). The groups did not differ by age (*p*=.40) nor by sex (*p*>.99). Lower scores were found on full-scale IQ (*p*<.001), performance IQ (*p*<.001), and verbal IQ (*p*=.010) in the RASopathies group compared to TD. Relative to TD, the RASopathies group also had lower scores on age-corrected cognitive measures of working memory, attention, receptive language, executive function, and expressive language (all *p*<.001). A power calculation to determine the minimum detectable effect sizes is available in the **Supplement.** Example QT1 images are shown in **Figure 2**.

**Figure 1.**
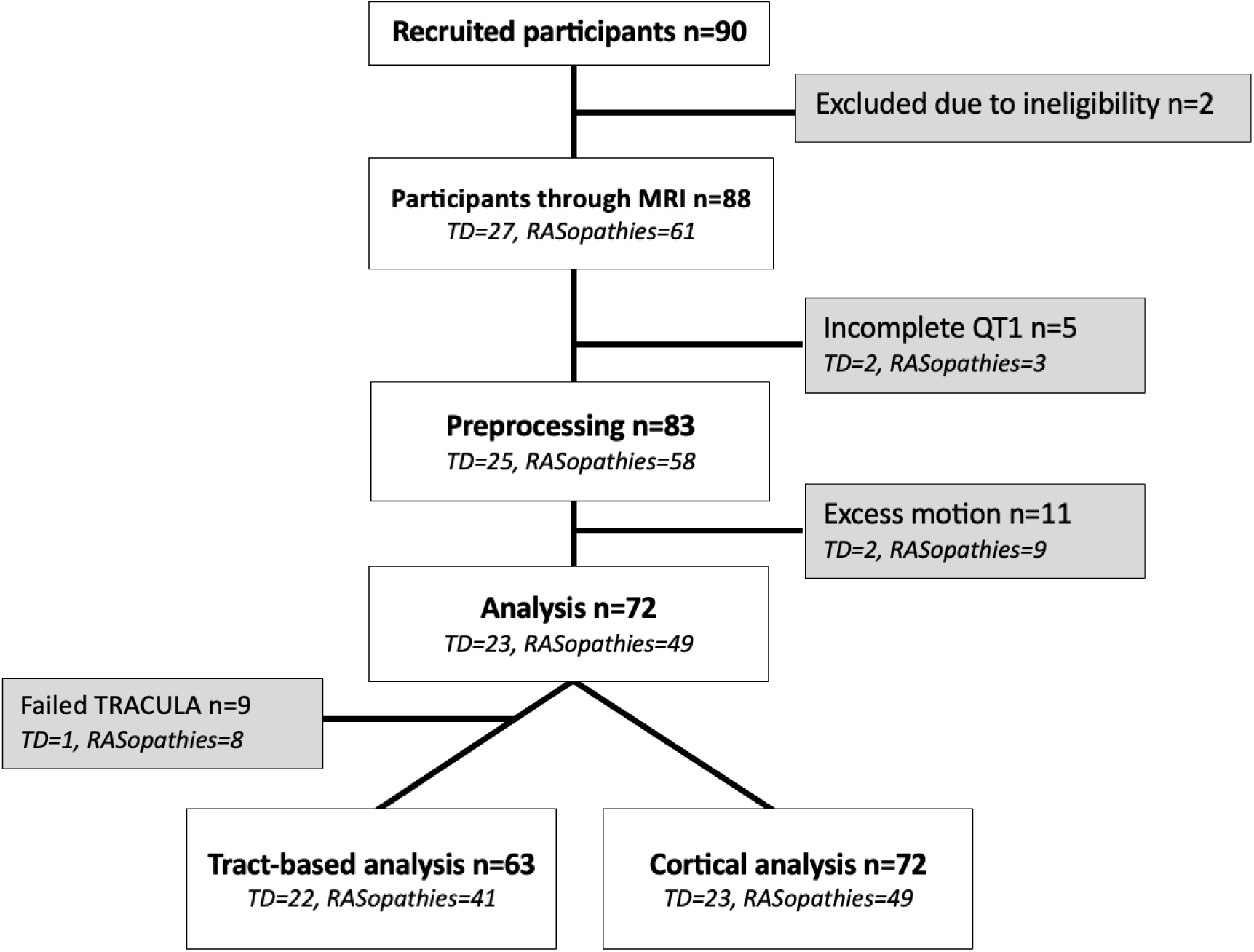
Flow of participants through the study. Following exclusions and quality checking, 72 subjects were included in cortical analysis and 65 subjects were included in tract-based analysis. *QT1=quantitative T1 mapping; TD=typical developing; TRACULA=TRActs Constrained by UnderLying Anatomy*.

**Figure 2.**
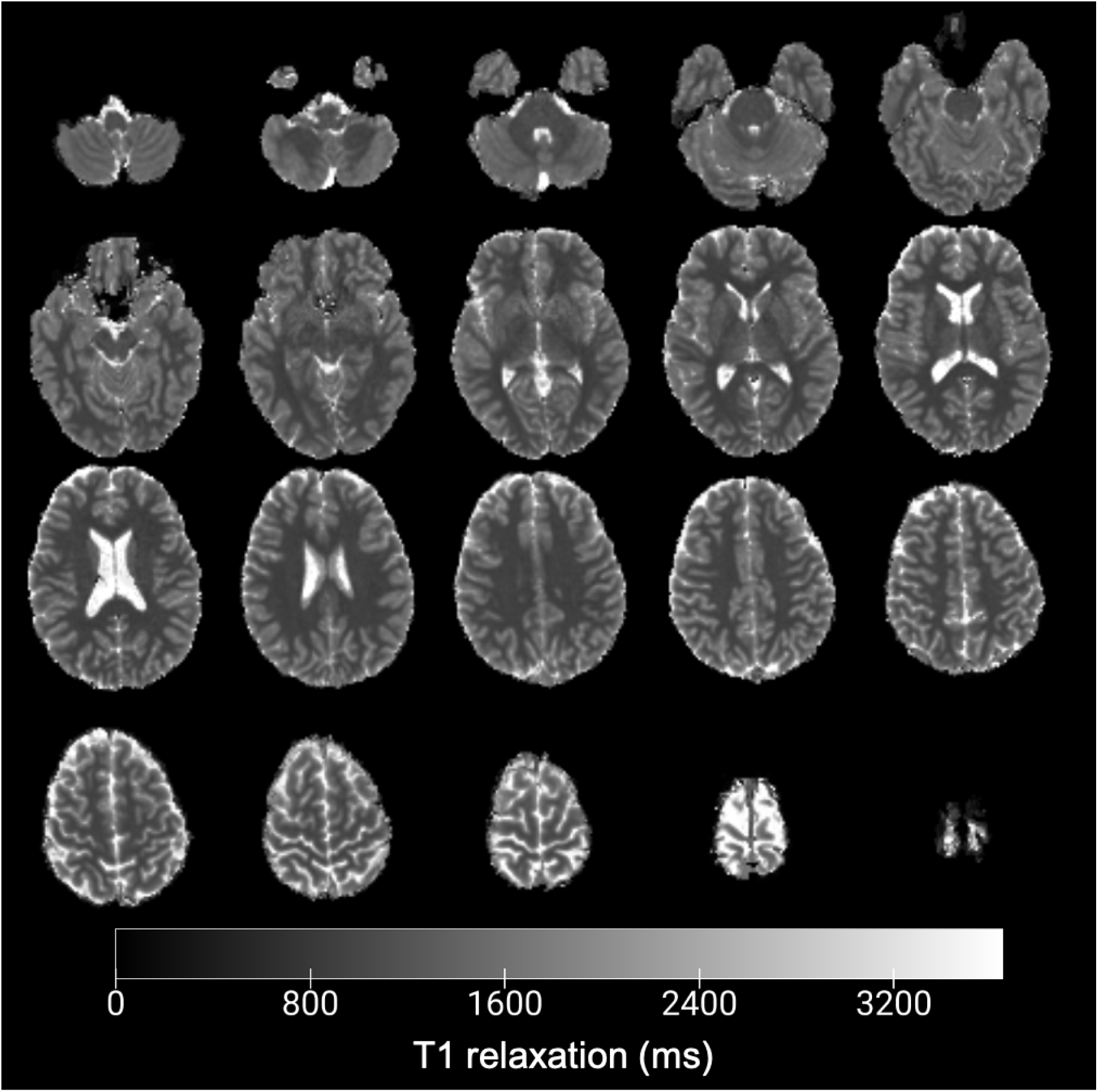
Example QT1 image slices from a single participant. Image created in MRICroGL.

**Table 1.** Demographics and descriptive statistics of included participants.

| | TD (n=23) | RAS (n=49) | $F/\chi^2(df)$ | $p$ |
| --- | --- | --- | --- | --- |
| **Age** | 10.2 (3.48) | 11.0 (3.48) | $F(1,70) = .729$ | .396 |
| **Sex (M/F)** | 11/12 | 25/24 | $\chi^2(1) = .000$ | >.999 |
| **Full-scale IQ** | 119 (14.7) | 96.3 (14.7) | $F(1,68) = 36.7$ | <.001 |
| **Verbal IQ** | 114 (19.8) | 101 (19.8) | $F(1,68) = 7.04$ | .010 |
| **Performance IQ** | 120 (13.7) | 94.4 (16.0) | $F(1,68) = 42.9$ | <.001 |
| **Working memory<sup>†</sup>** | 107 (14.2) | 92.7 (15.1) | $F(1,69) = 14.8$ | <.001 |
| **Attention<sup>†</sup>** | 96.3 (14.2) | 84.0 (10.1) | $F(1,69) = 18.3$ | <.001 |
| **Receptive language<sup>†</sup>** | 118 (17.6) | 100 (14.5) | $F(1,69) = 20.6$ | <.001 |
| **Episodic memory<sup>†</sup>** | 106 (17.0) | 100 (16.0) | $F(1,69) = 1.80$ | .184 |
| **Executive function<sup>†</sup>** | 111 (15.8) | 90.6 (15.0) | $F(1,68) = 29.0$ | <.001 |
| **Expressive language<sup>†</sup>** | 111 (15.1) | 91.9 (12.2) | $F(1,69) = 31.0$ | <.001 |
| **Processing speed<sup>†</sup>** | 91.4 (21.2) | 82.2 (19.9) | $F(1,68) = 3.10$ | .083 |

Data presented as counts or mean (SD). Between-group differences were calculated using analysis of covariance, including covariates of age and sex, or Chi-square test. Presented $p$ -values are uncorrected. Two subjects (both from RASopathies group) did not complete executive function and processing speed measures. <sup>†</sup>age-corrected scores derived from NIH toolbox tests: working memory (List Sorting Working Memory Test), attention (Flanker Inhibitory Control and Attention Test), receptive language (Picture Vocabulary Test), episodic memory (Picture Sequence Memory Test), executive function (Dimensional Change Card Sort Test), expressive language (Oral Reading Recognition Test), and processing speed (Pattern Comparison Processing Speed Test).
RAS = RASopathies; TD= typical developing.

### Children with RASopathies demonstrate higher MTV, suggesting greater myelin content, in majority of white matter tracts

Nine subjects were excluded from tract-based analysis (total n=63) due to motion in the diffusion-weighted images and subsequent failure to form tracts in TRACULA (**Figure 1**). The anterior commissure, left fornix, and right fornix were excluded from all tract-based analysis due to a low number of subjects with acceptable reconstructions (<80% acceptable). Following removal of these three tracts, 1.67% of all tract-based data were missing. The missing values were subsequently imputed using a predictive mean matching algorithm *Multiple Imputation by Chained Equations (MICE)* package in R.

Thirty-four out of 39 white matter tracts investigated showed significant between-group differences (*p_FDR_*<.05). Higher MTV was found in the RASopathies group relative to TD, indicating greater white matter myelin in RASopathies (**Figure 3**). We found the largest effect sizes (largest absolute value of Cohen’s *d*) in the right superior longitudinal fasciculus II (*p_FDR_*=.006, *d*=1.01), right anterior thalamic radiation (*p_FDR_*=.007, *d*=0.95), left cingulum bundle-dorsal (*p_FDR_*=.007, *d*=0.91), and corpus callosum-rostrum (*p_FDR_*=.007, *d*=0.86). The average MTV values in each group are shown in **Supplementary Table 1**.

**Figure 3.**
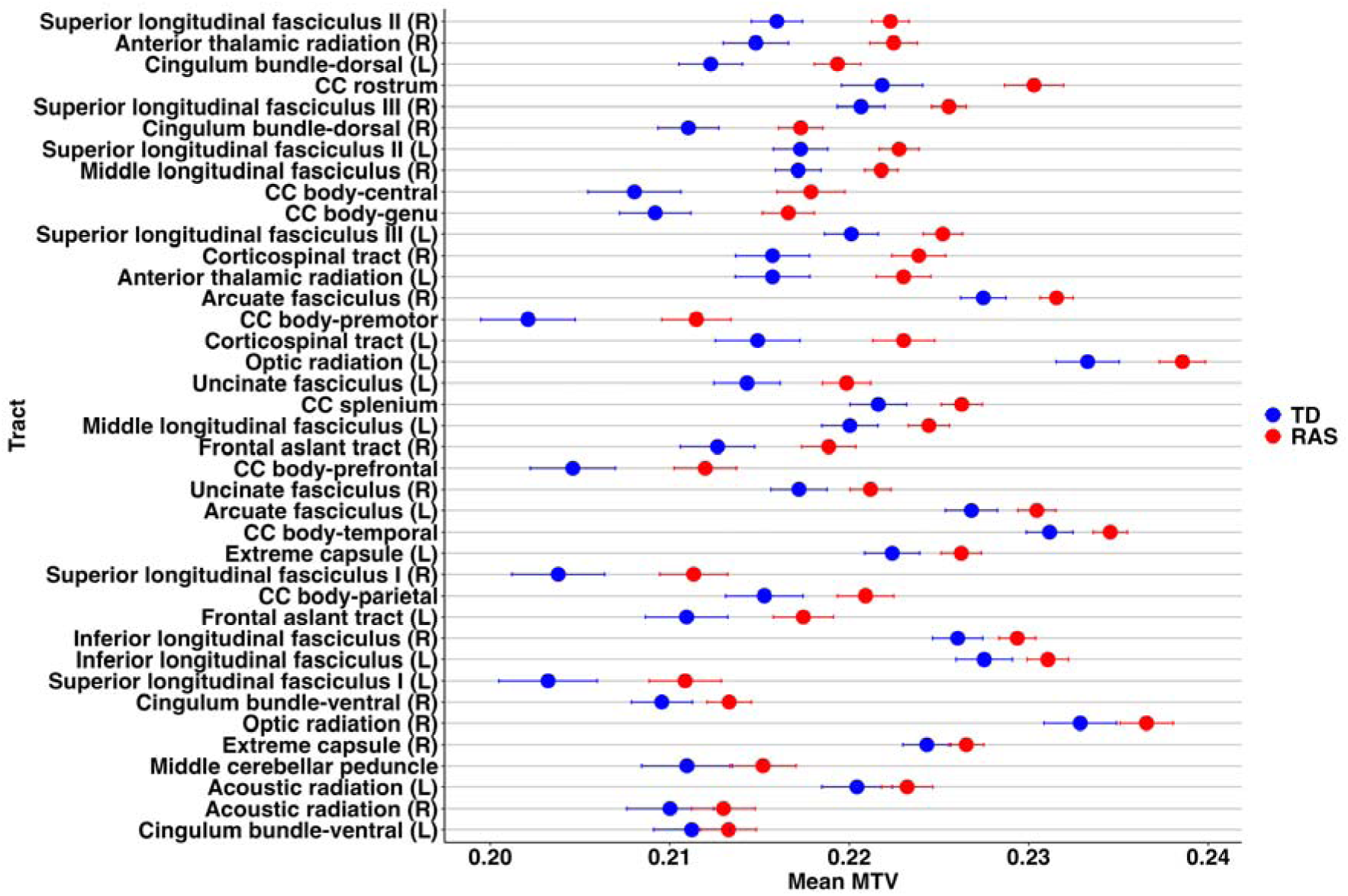
Average tract MTV and standard errors for each group (TD, RAS), ordered from largest effect size at the top to smallest effect size at the bottom. Thirty-four of 39 tracts had significantly elevated MTV in the RAS group relative to TD (*p_FDR_*<.05), suggesting greater white matter myelin content in the RAS group. *CC=corpus callosum; L=left; MTV=macromolecular tissue volume; R=right; RAS=RASopathies; TD=typical developing*.

We examined the two groups of RASopathies, Noonan syndrome (n=29) and neurofibromatosis type 1 (n=12), separately to gain a deeper understanding of the specific myelination patterns associated with each disorder. However, we did not find any differences in tract MTV between Noonan syndrome and neurofibromatosis type 1, indicating similar tract myelin in both groups. Statistical comparisons between TD and Noonan syndrome revealed 17 out of 39 tracts with higher MTV (*p_FDR_*<.05, **Figure 4A**), whereas comparisons between TD and neurofibromatosis type 1 revealed 36 of 39 tracts with higher MTV relative to TD (*p_FDR_*<.05, **Figure 4B**). Overall, both RASopathies showed elevations in tract MTV, suggesting greater myelin, relative to TD.

**Figure 4.**
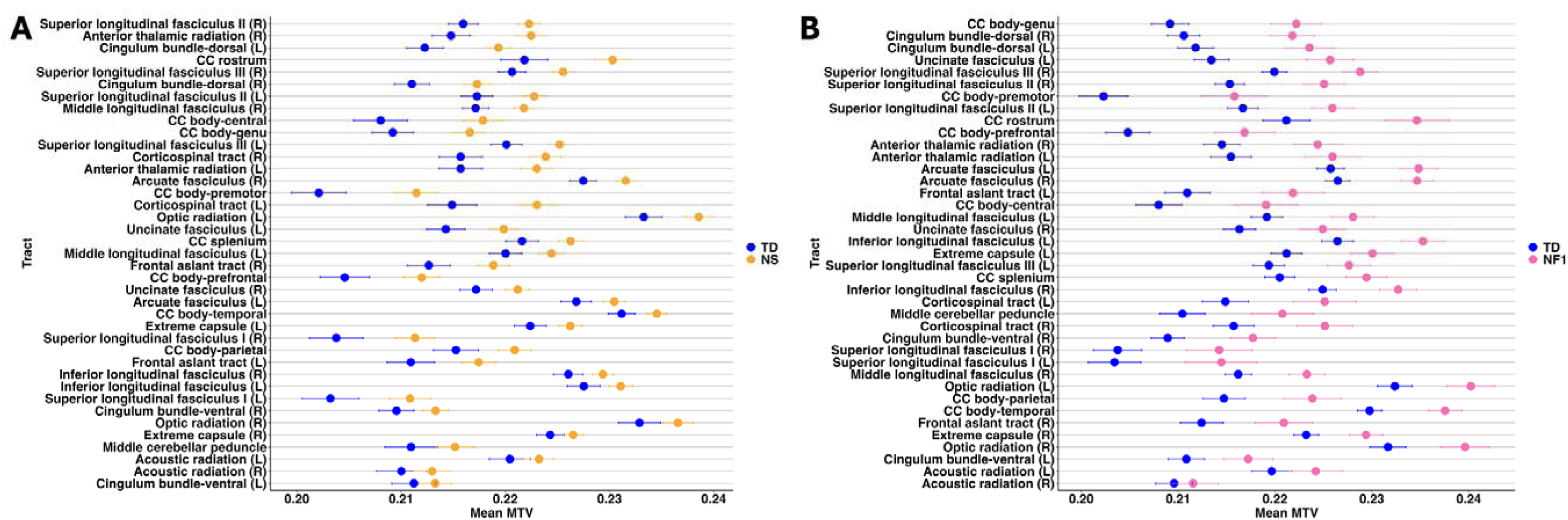
Average tract MTV and standard errors in each of the RASopathies compared to TD, ordered from largest effect size at the top to smallest effect size at the bottom. A) Seventeen of 39 tracts have significantly elevated MTV in Noonan syndrome relative to TD (*p_FDR_*<.05). B) Thirty-six of 39 tracts had significantly elevated MTV in neurofibromatosis type 1 relative to TD (*p_FDR_*<.05). Overall, the results indicate greater white matter myelin in both groups relative to TD. *CC=corpus callosum; L=left; MTV=macromolecular tissue volume; NF1=neurofibromatosis type 1; NS=noonan syndrome; R=right; RAS=RASopathies; TD=typical developing*.

### Children with RASopathies tend to have longer R1 (greater myelin content) in regions adjacent to the hippocampal formation

We conducted a PCA followed by MANOVA to analyze R1 (units 1/s) across all 360 cortical regions. The PCA reduced the dimensionality of the 360 regions to 71 principal components. Calculation of the Mahalanobis distance revealed zero outlier subjects at a threshold for exclusion of *p*<.001. The first and second principal components (PCs) individually explained 18% and 5.3% of the variance, respectively. Twenty-two PCs, explaining 67% of the cumulative variance, were entered as dependent variables into a non-parametric MANOVA with group (TD, RASopathies) as the independent variable. A scree plot shows the variance explained by each of the 22 PCs (**Supplementary Figure 1**). The MANOVA showed an effect of group on the cortical R1 PCs (Wilks’ Lambda=0.54, *F*(22,70)=2.73, *p*=.002, *η^2^*=.028). Visualization of the top PCs suggests subjects with RASopathies tend to have higher scores on PC2 relative to TD (**Figure 5A, 5B**), which likely contributed to the significant effect detected by the MANOVA. The ten regions with the greatest contributions to PC2 (**Table 2**) include several regions within the hippocampal area of the brain including R1 of the right PreSubiculum, right ParaHippocampal Area 3, and bilateral Entorhinal Cortex. The results suggest subjects with RASopathies tend to have longer R1 (greater myelin content) in these regions compared to TD.

**Figure 5.**
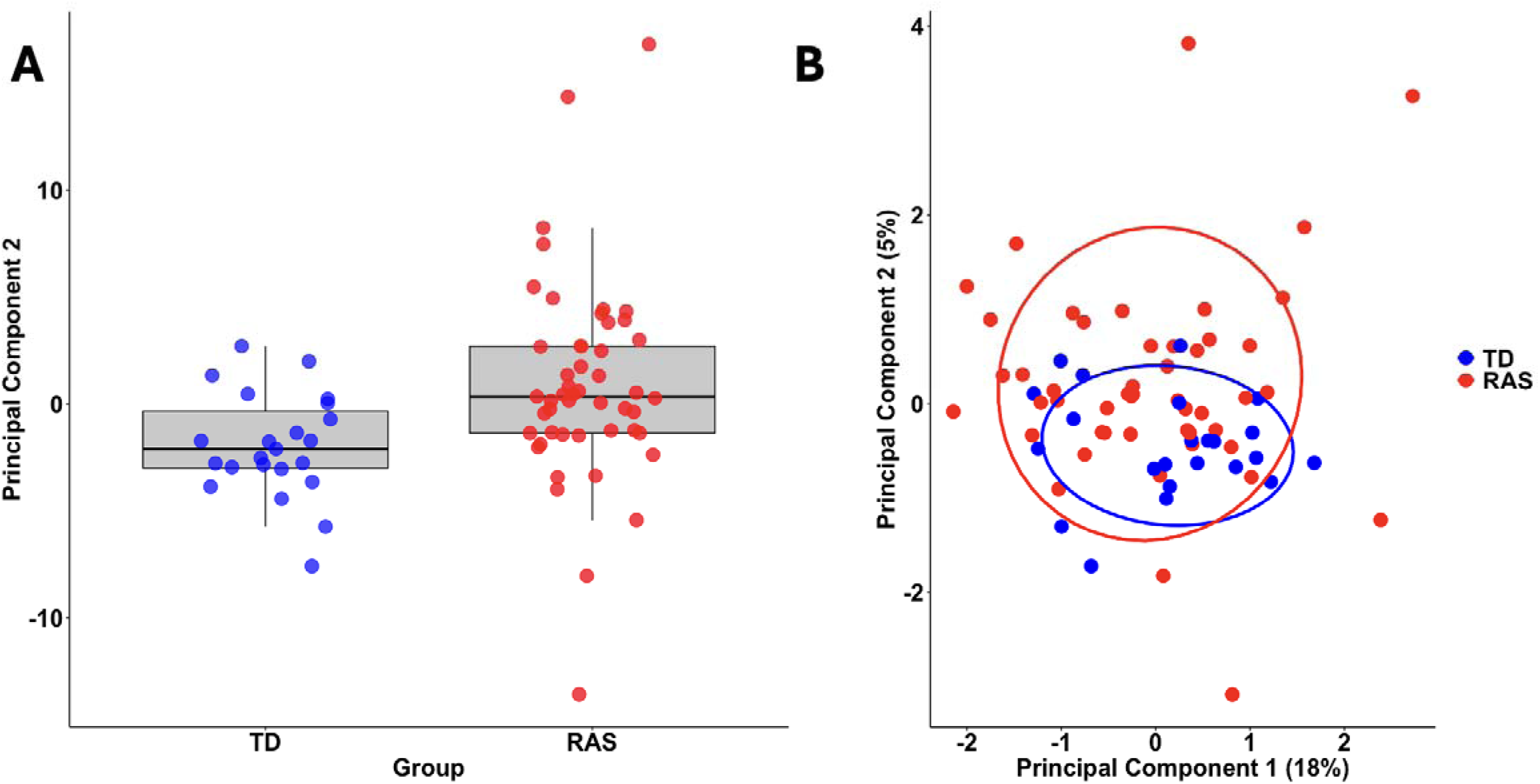
Subjects with RASopathies tend to have higher scores on PC2 relative to TD. (A) Boxplot visualizations suggest that subjects with RASopathies (pink) score higher on PC2 than TD subjects (blue). A Wilcoxon rank-rum test indicates the difference is significant *Z*=3.38, *p*=.008, *r*=0.398. (B) A biplot of PC 1 and PC2 indicates that TD subjects tend to score similarly on both PC1 and PC2, whereas subjects with RASopathies show more variation in scores on both PCs.

**Table 2.** The ten regions with the greatest weightings in each of PC1 and PC2.

| PC1 |  | PC2 |  |
| --- | --- | --- | --- |
| Region | Weighting | Region | Weighting |
| Area Lateral IntraParietal dorsal (R) | 0.51 | PreSubiculum (R) | 0.39 |
| Area Lateral Occipital 2 (R) | 0.51 | Area PH (L) | 0.36 |
| Auditory 4 Complex (L) | 0.51 | Entorhinal Cortex (L) | 0.32 |
| PreCuneus Visual Area (L) | 0.48 | Area 8B Lateral (L) | 0.31 |
| Area 8C (R) | 0.47 | PeriSylvian Language Area (L) | 0.31 |
| ParaHippocampal Area 1 (R) | 0.47 | Inferior 6-8 Transitional Area (L) | 0.31 |
| Sixth Visual Area (L) | 0.44 | ParaHippocampal Area 3 (R) | 0.29 |
| Area 44 (R) | 0.43 | Entorhinal Cortex (R) | 0.28 |
| Area TemporoParietoOccipital Junction 2 (R) | 0.42 | Frontal Opercular Area 3 (L) | 0.27 |
| Area 45 (L) | 0.41 | Primary Motor Cortex (R) | 0.26 |

L=left; PC=principal component; R=right.

### Support vector machine (SVM) suggests select cognitive and cortical R1 features are key discriminants between RASopathies and TD

We used SVM to determine the most discriminant features between children with RASopathies and TD. Tract MTV, cortical R1, and the cognitive measures were used to produce the binary SVM classifier (RASopathies versus TD). Sixty-one subjects were included in the SVM. Two subjects were excluded following tract-based analysis (n=63) due to missing values in their cognitive data. The accuracy, sensitivity, and specificity of the classifier were 86.9%, 86.4%, and 87.2%, respectively. The ROC curve is shown in **Figure 6A**. The area under the curve (AUC) is 0.95. The *p*-values from the SVM permutation-testing comparison demonstrated that the accuracy of the SVM was significantly greater than chance (*p*<.001), i.e., the SVM has a significantly higher accuracy (87%) than the null model (57% average accuracy).

**Figure 6.**
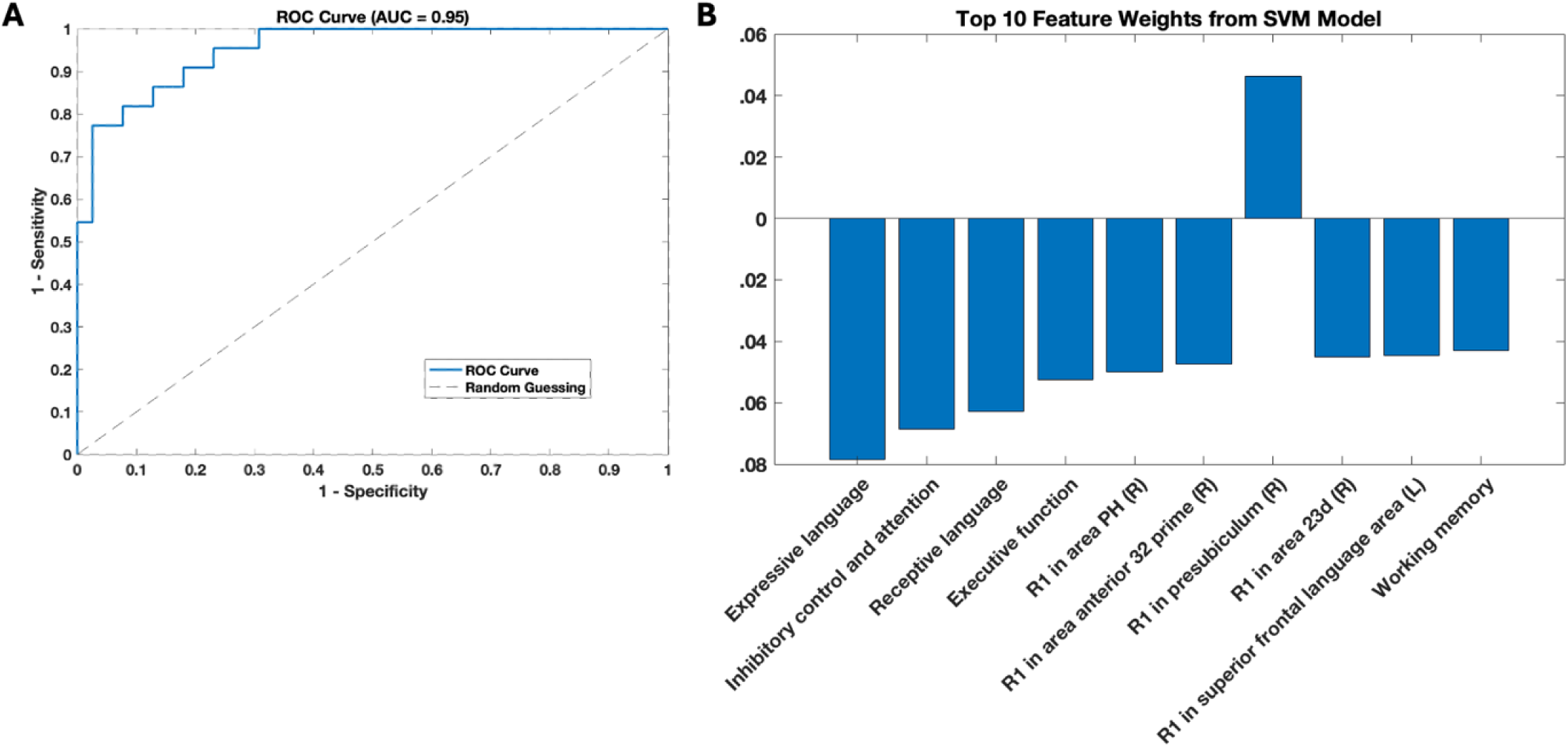
SVM Classifier. A) ROC Curve, area under the curve (AUC) is 0.95. B) Top 10 feature weights from the SVM classifier. Expressive language (Oral Reading Recognition Test), inhibitory control and attention (Flanker Inhibitory Control and Attention Test), receptive language (Picture Vocabulary Test), executive function (Dimensional Change Card Sort Test), and working memory (List Sorting Working Memory Test) are derived from the NIH toolbox tests named in brackets. *L=left; R=right; ROC=receiver operating characteristic*

The ten features with the greatest weights are shown in **Table 3 and Figure 6B**. The positive weights suggest subjects with increases in these features are more likely to be classified as RASopathies, whereas the negative weights indicate increases in these features are more likely to be classified as TD. Our results suggest longer R1 values in the right presubiculum, a region located in the temporal lobe between the hippocampus and entorhinal cortex, mean the subject is more likely to be classified in the RASopathies class. The nine remaining features out of the top ten were all negative, suggesting increases in these features mean the subject is more likely to be classified as TD. Five of these features were cognitive measures including language, inhibitory control and attention, executive function, and working memory. The remaining four features were R1 measurements from cortical regions including regions located in the superior frontal gyrus (right superior frontal language area and right area anterior 32 prime), area PH located between the temporal and occipital lobes, and area 23d in the posterior cingulate.

**Table 3.** Ten features with the greatest weights in the classifier generated by SVM.

| Feature | Weight |
| --- | --- |
| Expressive language | -0.078 |
| Inhibitory control and attention | -0.069 |
| Receptive language | -0.063 |
| Executive function | -0.053 |
| R1 in Area PH (R) | -0.050 |
| R1 in Area anterior 32 prime (R) | -0.047 |
| R1 in PreSubiculum (R) | 0.046 |
| R1 in Area 23d (R) | -0.045 |
| R1 in Superior frontal language area (L) | -0.045 |
| Working memory | -0.043 |
| Expressive language (Oral Reading Recognition Test), inhibitory control and attention (Flanker Inhibitory Control and Attention Test), executive function (Dimensional Change Card Sort Test), receptive language (Picture Vocabulary Test), and working memory (List Sorting Working Memory Test) are derived from the NIH toolbox tests named in brackets. |  |

Five subjects with RASopathies (mutations of *PTPN11*=2*, SOS-1*=2*, RAF-1*=1) were incorrectly classified as TD in the SVM following the leave-one-out cross-validation. The values of the top 10 features for each of these five incorrectly classified subjects are shown in **Supplementary Table 2**. Compared to the correctly classified RASopathies subjects, all five of the incorrectly classified subjects had higher inhibitory control and attention scores (83, 88, 100, 101, and 84) compared to the group average (81). These subjects also had longer R1 of the left superior frontal language area (0.618, 0.594, 0.629, 0.617, 0.641) compared to the group average (0.586) of the correctly classified RASopathies subjects.

Three TD subjects were incorrectly classified as RASopathies. The values of the top 10 features for each of these TD subjects are shown in **Supplementary Table 3**. The incorrectly classified subjects had lower expressive language scores (100, 89, 90) and inhibitory control and attention scores (92, 90, 83) compared to the group averages of the correctly classified TD subjects (114 and 99, respectively). The subjects also had longer R1 of the right presubiculum (0.606, 0.613, 0.615) compared to the group average (0.602).

As five of the most discriminant ten features in the SVM were cognitive measures, we were interested in investigating the added value of brain-based measures. We tested the difference in classification performance between the two models by running a second SVM using seven cognitive measures but excluding any brain-based metrics. The SVM classifier based on cognitive measures alone yielded accuracy, sensitivity, and specificity of 77.1%, 72.7%, and 79.5%, respectively. The ROC curve is shown in **Supplementary Figure 2**. The area under the curve (AUC) is 0.89. Fifteen subjects (RASopathies=8, TD=6) were classified incorrectly following the leave-one-out cross-validation. Overall, these findings indicate that integrating brain-based with cognitive measures enhances the accuracy of the SVM by 10%, emphasizing the added value of a multimodal approach for understanding RASopathies.

## Discussion

Using QT1 mapping, we identified differences in white matter tracts and cortical myelin content in children with RASopathies relative to TD peers. Our analysis of tract MTV revealed widespread elevations in white matter myelin in RASopathies, to the extent that thirty-four out of thirty-nine (87%) showed significantly increased MTV compared to TD. The results of our MANOVA suggest group differences in cortical myelination, with a significant effect driven partially by PC2. Higher scores on PC2 were found in the RASopathies group relative to TD, suggesting children with RASopathies may have longer R1 (increased myelin) of the contributing regions. Finally, the SVM identified cognitive and neuroimaging-based features that discriminate the RASopathies and TD groups with reasonable accuracy. The most discriminating features indicate the strong effects of RASopathies on language and cognitive abilities, and on cortical myelination. Taken together, our findings illustrate the extensive impact of the RAS-MAPK mutations on myelination and demonstrate the potential of QT1 as a non-invasive measure of pathology in clinical populations.

Our analysis of tract MTV reveals increased myelin content in many white matter tracts in children with RASopathies compared to their TD peers. Prior *in vivo* studies of RASopathies using DTI have identified white matter alterations, such as reduced FA and increased mean diffusivity (MD), in children with neurofibromatosis type 1 and Noonan syndrome (13,14,16,17). While these findings suggest abnormalities in white matter structure, FA and MD are non-specific metrics influenced by multiple factors, including myelin content, axonal density, and fiber orientation coherence (9,10). For example, FA may reflect both the degree of myelination and alignment of axons within a voxel, making it difficult to isolate the specific biological mechanisms. In contrast, QT1 measures like R1 provide more direct and specific information about myelin content. Unlike DTI, QT1 metrics are largely independent of fiber orientation and thus offer a clearer representation of myelin concentration (27). Supporting this, previous work demonstrated a weak relationship between R1 and diffusion measures in white matter, highlighting that these modalities capture distinct biological processes (12). By leveraging QT1 to measure tract MTV, our study advances understanding of RASopathies by identifying significant myelin increases without relying on the assumptions required by DTI. QT1 highlights specific myelin-related changes, providing novel insights into the white matter abnormalities associated with RASopathies.

To elucidate the mechanisms driving increased myelin content in RASopathies, we can leverage insights from animal models of these conditions. Animal models of neurofibromatosis type 1 and Noonan syndrome demonstrated proliferation of oligodendrocyte precursor cells (OPC), indicating alterations in the early stages of oligodendrocyte development (5,28,29). However, the impact on later processes involving the differentiation and functionality of mature oligodendrocytes remains unclear (30). For instance, a mouse model of Noonan syndrome showed fewer myelinated axons in white matter, leading to the hypothesis of hypomyelination (5). In contrast, our findings suggest white matter hypermyelination among children with RASopathies. Our findings are at least partially driven by the increased myelin detected in neurofibromatosis type 1, where 36 of 39 tracts were significantly higher in tract MTV relative to TD. Animals models of neurofibromatosis type 1 showed increased myelin thickness (31), hypermyelination, and myelin decompaction (32). Future longitudinal studies would be valuable to further evaluate the impact of the RAS-MAPK mutations on myelination trajectories *in vivo*.

Using multivariate techniques, including PCA and SVM classification, we found region-dependent differences in cortical myelin content within the RASopathies group relative to TD individuals. The PCA and MANOVA revealed several cortical regions adjacent to the hippocampal formation where myelin content is likely elevated in children with RASopathies. Interestingly, the hippocampus is involved in memory and working memory is often impaired in children with RASopathies (6), as we also found in the present study. In contrast, the SVM classifier detected multiple cortical regions where myelin content appeared lower in this group. Collectively, these findings suggest complex and varied effects of RAS-MAPK mutations on cortical myelination. These findings may also be related to atypical developmental trajectories observed in RASopathies compared to TD, characterized by certain regions that mature more quickly while others lag in development. The regions highlighted in the SVM analysis play a role in visuospatial processing – a cognitive domain affected in children with neurofibromatosis type 1 and Noonan syndrome (33,34). Further research incorporating functional MRI would be valuable to understand the potential link between the myelination of these regions and visuospatial processing in RASopathies.Additionally, our analysis revealed five cognitive measures that served as the most discriminative factors between TD individuals and those with RASopathies, aligning with previous studies that indicate lower cognitive scores in children with RASopathies compared to their TD peers (6,35–37). Importantly, however, the SVM based on cognitive measures alone yielded lower accuracy (77%) than the SVM based on cognitive and brain-based measures (87% accuracy), showing the value of multimodal approaches for improved precision. Ultimately, these findings enhance our understanding of myelination patterns in RASopathies while affirming the need for targeted interventions to support cognitive development in this population.

While this study was limited by small group sizes, we nonetheless focused on biologically defined neurodevelopmental disorders and found large effect sizes highlighting the clinical significance of this work. To our knowledge, this is the first study to apply QT1 mapping in children with RASopathies and our results suggest this technique is promising for clinical applications. Due to its relatively short scan time and specificity, QT1 may be instrumental in identifying objective treatment targets and subsequent monitoring of treatment efficacy over time. In our analysis we utilized a leave-one-out cross-validation to fit the SVM classifier. We acknowledge that a train-test split across 1000 iterations would provide a more rigorous validation process; however, the small sample size in each group was not conducive to such an approach. Future studies with larger sample sizes and more statistical power will be essential to validate and improve the robustness of these findings.

This study builds on prior research by utilizing QT1 mapping to examine changes in myelin content in children with RASopathies relative to TD peers. The evidence suggests widespread elevated myelin content in white matter tracts and region-dependent patterns of cortical myelination in children with RASopathies. The SVM suggests a unique pattern of myelin content in RASopathies that may be of relevance to visuospatial abilities. The significance of these findings warrants further investigation, ideally involving larger sample sizes to garner a more comprehensive and nuanced picture of the underlying neurobiology associated with the RAS-MAPK pathway. QT1 mapping demonstrates promise for clinical translation and should be explored further in additional RASopathies and neurodevelopmental disorders to ultimately develop novel treatments and improve outcomes for affected children.

## Data Availability

All data produced in the present study are available upon reasonable request to the authors.

## Acknowledgments

We thank the families who participated in this research. The authors would also like to thank the Noonan Syndrome Foundation, the RASopathies Network, and the Children’s Tumor Foundation which made this work possible. We would like to thank Stanford University and the Stanford Research Computing Center for providing computational resources and support that contributed to these research results, some of the computing for this project was performed on the Sherlock cluster. We gratefully acknowledge the support of The Lucas Service Center at Stanford.

## References

1. Dietrich KN, Eskenazi B, Schantz S, Yolton K, Rauh VA, Johnson CB, et al. Principles and practices of neurodevelopmental assessment in children: lessons learned from the Centers for Children’s Environmental Health and Disease Prevention Research. Environ Health Perspect. 2005 Oct;113(10):1437–46.

2. Zhao Y, Yang L, Gong G, Cao Q, Liu J. Identify aberrant white matter microstructure in ASD, ADHD and other neurodevelopmental disorders: A meta-analysis of diffusion tensor imaging studies. Prog Neuropsychopharmacol Biol Psychiatry. 2022 Mar 8;113:110477.

3. de Blank P, Nishiyama A, López-Juárez A. A new era for myelin research in Neurofibromatosis type 1. Glia. 2023 Dec;71(12):2701–19.

4. Buyanova IS, Arsalidou M. Cerebral White Matter Myelination and Relations to Age, Gender, and Cognition: A Selective Review. Front Hum Neurosci. 2021 Jul 6;15:662031.

5. Ehrman LA, Nardini D, Ehrman S, Rizvi TA, Gulick J, Krenz M, et al. The protein tyrosine phosphatase Shp2 is required for the generation of oligodendrocyte progenitor cells and myelination in the mouse telencephalon. J Neurosci. 2014 Mar 5;34(10):3767– 78.

6. Naylor PE, Bruno JL, Shrestha SB, Friedman M, Jo B, Reiss AL, et al. Neuropsychiatric phenotypes in children with Noonan syndrome. Dev Med Child Neurol. 2023 Nov;65(11):1520–9.

7. Pierpont EI, Tworog-Dube E, Roberts AE. Attention skills and executive functioning in children with Noonan syndrome and their unaffected siblings. Dev Med Child Neurol. 2015 Apr;57(4):385–92.

8. Elia J, Ungal G, Kao C, Ambrosini A, De Jesus-Rosario N, Larsen L, et al. Fasoracetam in adolescents with ADHD and glutamatergic gene network variants disrupting mGluR neurotransmitter signaling. Nat Commun. 2018 Jan 16;9(1):4.

9. Pierpaoli C, Jezzard P, Basser PJ, Barnett A, Di Chiro G. Diffusion tensor MR imaging of the human brain. Radiology. 1996 Dec;201(3):637–48.

10. Timmers I, Roebroeck A, Bastiani M, Jansma B, Rubio-Gozalbo E, Zhang H. Assessing Microstructural Substrates of White Matter Abnormalities: A Comparative Study Using DTI and NODDI. PLoS One. 2016 Dec 21;11(12):e0167884.

11. Mezer A, Yeatman JD, Stikov N, Kay KN, Cho N-J, Dougherty RF, et al. Quantifying the local tissue volume and composition in individual brains with magnetic resonance imaging. Nat Med. 2013 Dec;19(12):1667–72.

12. Yeatman JD, Wandell BA, Mezer AA. Lifespan maturation and degeneration of human brain white matter. Nat Commun [Internet]. 2014; Available from: https://www.nature.com/articles/ncomms5932

13. Fattah M, Raman MM, Reiss AL, Green T. PTPN11 Mutations in the Ras-MAPK Signaling Pathway Affect Human White Matter Microstructure. Cereb Cortex. 2021 Feb 5;31(3):1489–99.

14. Siqueiros-Sanchez M, Dai E, McGhee CA, McNab JA, Raman MM, Green T. Impact of pathogenic variants of the Ras-Mitogen-activated protein kinase (Ras-MAPK) pathway on major white matter tracts in the human brain. Brain Commun. 2024 Aug 14;6(4):fcae274.

15. Plank J, Gozdas E, Dai E, McGhee C, Raman M, Green T. Elucidating microstructural alterations in neurodevelopmental disorders: application of advanced diffusion-weighted imaging in children with Rasopathies. Human Brain Mapping. 2024 In Press; Available from: https://www.researchsquare.com/article/rs-4415218/latest

16. Braun-Walicka N, Pluta A, Wolak T, Maj E, Maryniak A, Gos M, et al. Research on the pathogenesis of cognitive and neurofunctional impairments in patients with Noonan syndrome: The role of rat sarcoma–mitogen activated protein kinase signaling pathway gene disturbances. Genes [Internet]. 2023 Dec 1;14. Available from: https://www.mdpi.com/2073-4425/14/12/2173

17. Ferraz-Filho JRL, da Rocha AJ, Muniz MP, Souza AS, Goloni-Bertollo EM, Pavarino-Bertelli EC. Diffusion tensor MR imaging in neurofibromatosis type 1: expanding the knowledge of microstructural brain abnormalities. Pediatr Radiol. 2012 Apr;42(4):449– 54.

18. Wechsler D. Wechsler Abbreviated Scale of Intelligence [Internet]. 1999. Available from: https://psycnet.apa.org/fulltext/9999-15170-000.pdf

19. Gershon RC, Wagster MV, Hendrie HC, Fox NA, Cook KF, Nowinski CJ. NIH Toolbox for Assessment of Neurological and Behavioral Function. Neurology [Internet]. 2013 Mar 12;80(11_supplement_3). Available from: https://www.neurology.org/doi/abs/10.1212/wnl.0b013e3182872e5f?casa_token=esdRkwLn8ckAAAAA:LYbyIq0Yl52AVaG5upZZVk1OBey8gHVfYeLUWOB7nd1FkPyP28AuboZzXPhAdQgqiVYj9ZKsuFO8CJHA&casa_token=WpqJyeb_NzEAAAAA:oNIUKQalh59g-9oSEzhXazhoMb5khmJ5QTqAPOTNA7Jve4pbPGdln1_IViy59ydhiuoKBnhkchKwrGYn

20. Wu H, Dougherty R, Kerr AB, Zhu K, Middione MJ, Mezer A. Fast T1 mapping using slice-shuffled Simultaneous Multi-Slice inversion recovery EPI. In: 21st Annual Meeting of the Organization for Human Brain Mapping [Internet]. archive.ismrm.org; 2015. Available from: https://archive.ismrm.org/2015/0440.html

21. Andersson JLR, Skare S, Ashburner J. How to correct susceptibility distortions in spin-echo echo-planar images: application to diffusion tensor imaging. Neuroimage. 2003 Oct;20(2):870–88.

22. Andersson JLR, Sotiropoulos SN. An integrated approach to correction for off-resonance effects and subject movement in diffusion MR imaging. Neuroimage. 2016 Jan 15;125:1063–78.

23. Glasser MF, Coalson TS, Robinson EC, Hacker CD, Harwell J, Yacoub E, et al. A multi-modal parcellation of human cerebral cortex. Nature. 2016 Aug 11;536(7615):171–8.

24. Gozdas E, Fingerhut H, Wu H, Bruno JL, Dacorro L, Jo B, et al. Quantitative measurement of macromolecular tissue properties in white and gray matter in healthy aging and amnestic MCI. Neuroimage. 2021 Aug 15;237:118161.

25. Yendiki A, Panneck P, Srinivasan P, Stevens A, Zöllei L, Augustinack J, et al. Automated probabilistic reconstruction of white-matter pathways in health and disease using an atlas of the underlying anatomy. Front Neuroinform. 2011 Oct 14;5:23.

26. Burchett WW, Ellis AR, Harrar SW, Bathke AC. Nonparametric inference for multivariate data: The R package npmv. J Stat Softw. 2017;76(4):1–18.

27. Stüber C, Morawski M, Schäfer A, Labadie C, Wähnert M, Leuze C, et al. Myelin and iron concentration in the human brain: a quantitative study of MRI contrast. Neuroimage. 2014 Jun;93 Pt 1:95–106.

28. Bennett MR, Rizvi TA, Karyala S, McKinnon RD, Ratner N. Aberrant growth and differentiation of oligodendrocyte progenitors in neurofibromatosis type 1 mutants. J Neurosci. 2003 Aug 6;23(18):7207–17.

29. Holter MC, Hewitt LT, Koebele SV, Judd JM, Xing L, Bimonte-Nelson HA, et al. The Noonan Syndrome-linked Raf1L613V mutation drives increased glial number in the mouse cortex and enhanced learning. PLoS Genet. 2019 Apr;15(4):e1008108.

30. Gutmann DH, Parada LF, Silva AJ, Ratner N. Neurofibromatosis type 1: modeling CNS dysfunction. J Neurosci. 2012 Oct 10;32(41):14087–93.

31. López-Juárez A, Titus HE, Silbak SH, Pressler JW, Rizvi TA, Bogard M, et al. Oligodendrocyte Nf1 controls aberrant Notch activation and regulates myelin structure and behavior. Cell Rep. 2017 Apr 18;19(3):545–57.

32. Kim KY, Ju WK, Hegedus B, Gutmann DH, Ellisman MH. Ultrastructural characterization of the optic pathway in a mouse model of neurofibromatosis-1 optic glioma. Neuroscience. 2010 Sep 29;170(1):178–88.

33. Vogel AC, Gutmann DH, Morris SM. Neurodevelopmental disorders in children with neurofibromatosis type 1. Dev Med Child Neurol. 2017 Nov;59(11):1112–6.

34. Alfieri P, Cesarini L, De Rose P, Ricci D, Selicorni A, Menghini D, et al. Visual processing in Noonan syndrome: dorsal and ventral stream sensitivity. Am J Med Genet A. 2011 Oct;155A(10):2459–64.

35. Johnson EM, Ishak AD, Naylor PE, Stevenson DA, Reiss AL, Green T. PTPN11 Gain-of-Function Mutations Affect the Developing Human Brain, Memory, and Attention. Cereb Cortex. 2019 Jul 5;29(7):2915–23.

36. Pierpont EI, Ellis Weismer S, Roberts AE, Tworog-Dube E, Pierpont ME, Mendelsohn NJ, et al. The language phenotype of children and adolescents with Noonan syndrome. J Speech Lang Hear Res. 2010 Aug;53(4):917–32.

37. Lehtonen A, Howie E, Trump D, Huson SM. Behaviour in children with neurofibromatosis type 1: cognition, executive function, attention, emotion, and social competence. Dev Med Child Neurol. 2013 Feb;55(2):111–25.

