## Supplementary for "Quantitative T1 mapping indicates elevated white matter myelin in children with RASopathies"

**Supplementary Material**

Methods and Materials

**Inclusion criteria**

*RASopathies*

Proof of genetic testing was required for individuals with RASopathies to demonstrate the presence of appropriate mutations. The inclusion criteria for all participants with RASopathies are as follows: a) age from 5 years, 0 months to 17 years, 11 months; b) IQ criterion ≥ 70; and c) gestational age > 34 weeks. Participants with neurofibromatosis type 1 and focal areas of signal intensity with no mass effect and/or low-grade optic pathway glioma are not excluded.

*Typical developing (TD)*

TD participants are group-matched to participants with RASopathies based on age, sex, handedness, socioeconomic status, pubertal status, and ethnicity. Subjects are included if their full-scale IQ is greater than 80.

**Exclusion criteria**

For all participants, the exclusion criteria includes: a) presence of neurological or psychiatric disease (e.g., psychotic symptoms); b) sensory deficits that would preclude participation in assessments or imaging; c) history of significant head trauma with loss of consciousness; d) use of psychotropic medications or currently taking medications with central nervous system effects within 5 half-lives of the scan; e) contraindications to MRI (e.g., metal implants, orthodontia, claustrophobia); f) history of alcohol or drug use; g) premature birth (<34 weeks); h) low birth weight (<2000g); i) history of head trauma with loss of consciousness; neurological disorders known to affect cognitive development or brain structure; j) known presence of gliomas in cerebellum, brainstem, or basal ganglia; and k) contraindications to MRI.

Additionally, participants with neurofibromatosis type 1 were excluded if they received an MEK inhibitor or chemotherapy, and/or show presence of cerebellar, brainstem, tectal plate, and basal ganglia gliomas. Participants with RASopathies were not excluded if they show clinical symptoms associated with typical syndromic cognitive-behavioral features (e.g., inattention, anxiety, learning disorder, or autistic traits).

Pubertal status was determined using Tanner staging (1,2). A full-scale intelligence quotient (IQ) ≥ 70 was required to maximize study compliance. Full-scale, performance, and verbal IQs were acquired using the Wechsler Abbreviated Scale of Intelligence 2^nd^ Edition (3).

**Cognitive measures**

The NIH Toolbox Cognition Battery (<http://www.nihtoolbox.org/>) was used to administer seven tests of cognition: the Dimensional Change Card Sort Test (assesses executive function) (4), Flanker Inhibitory Control and Attention Test (executive function, attention) (5), List Sorting Working Memory Test (memory, working) (6,7), Oral Reading Recognition Test (language, expressive) (8,9), Pattern Comparison Processing Speed Test (processing speed) (10), Picture Sequence Memory Test (memory, episodic) (11), and the Picture Vocabulary Test (language, receptive) (8,9). The Dimensional Change Card Sort Test requires participants to match picture pairs to a target picture. The Flanker Inhibitory Control and Attention Test asks participants to focus on a target stimulus and simultaneously ignore flanker stimuli. In the Oral Reading Recognition Test, participants are presented with a series of words and asked to pronounce each one as accurately as possible. The Pattern Comparison Processing Speed Test requires participants to quickly determine whether two stimuli are identical or not. In the Picture Sequence Memory Test, participants are shown a series of activities with pictures and are then asked to reproduce the picture sequence in the identical order as presented to them. The Picture Vocabulary Test presents participants with four pictures and asks participants to select which picture best matches the meaning of a word presented via audio.

**Results**

**Power calculation**

Given the absence of prior work using quantitative T1 mapping in RASopathies, there are no previous studies on which to base our sample size justification. We collected a final sample for analysis of 49 subjects with RASopathies and 23 TD. At an α=0.05 and 1-β=0.8, the minimum detectable effect size is therefore *d*=0.718. When comparing subjects with Noonan syndrome (n=34) and TD, the minimum detectable effect size is *d*=0.770. When comparing subjects with neurofibromatosis type 1 (n=15) and TD, the minimum detectable effect size is *d*=0.956. When comparing subjects with Noonan syndrome and neurofibromatosis type 1, the minimum detectable effect size is *d*=0.887.

**Supplementary Table 1. Average tract MTV in each group (TD, RASopathies).**

|  | **TD** | | **RASopathies** | |  |  |
| --- | --- | --- | --- | --- | --- | --- |
| **Tract** | **Mean** | **SE** | **Mean** | **SE** | **Cohen’s *d*** | ***p_FDR_*** |
| **Acoustic radiation (L)** | 0.220 | 0.002 | 0.223 | 0.001 | -0.397 | 0.121 |
| **Acoustic radiation (R)** | 0.210 | 0.002 | 0.213 | 0.002 | -0.308 | 0.251 |
| **Anterior thalamic radiation (L)** | 0.216 | 0.002 | 0.223 | 0.002 | -0.788 | 0.004 |
| **Anterior thalamic radiation (R)** | 0.215 | 0.002 | 0.222 | 0.001 | -0.953 | 0.001 |
| **Arcuate fasciculus (L)** | 0.227 | 0.001 | 0.230 | 0.001 | -0.627 | 0.007 |
| **Arcuate fasciculus (R)** | 0.227 | 0.001 | 0.232 | 0.001 | -0.754 | 0.001 |
| **CC body-central** | 0.208 | 0.003 | 0.218 | 0.002 | -0.817 | 0.003 |
| **CC body-genu** | 0.209 | 0.002 | 0.217 | 0.001 | -0.817 | 0.003 |
| **CC body-parietal** | 0.215 | 0.002 | 0.221 | 0.002 | -0.616 | 0.023 |
| **CC body-prefrontal** | 0.205 | 0.002 | 0.212 | 0.002 | -0.652 | 0.017 |
| **CC body-premotor** | 0.202 | 0.003 | 0.211 | 0.002 | -0.744 | 0.007 |
| **CC body-temporal** | 0.231 | 0.001 | 0.235 | 0.001 | -0.625 | 0.002 |
| **CC rostrum** | 0.222 | 0.002 | 0.230 | 0.002 | -0.863 | 0.001 |
| **CC splenium** | 0.222 | 0.002 | 0.226 | 0.001 | -0.709 | 0.003 |
| **Cingulum bundle-dorsal (L)** | 0.212 | 0.002 | 0.219 | 0.001 | -0.907 | 0.001 |
| **Cingulum bundle-dorsal (R)** | 0.211 | 0.002 | 0.217 | 0.001 | -0.853 | 0.002 |
| **Cingulum bundle-ventral (L)** | 0.211 | 0.002 | 0.213 | 0.002 | -0.258 | 0.330 |
| **Cingulum bundle-ventral (R)** | 0.210 | 0.002 | 0.213 | 0.001 | -0.546 | 0.038 |
| **Corticospinal tract (L)** | 0.215 | 0.002 | 0.223 | 0.002 | -0.734 | 0.008 |
| **Corticospinal tract (R)** | 0.216 | 0.002 | 0.224 | 0.002 | -0.803 | 0.003 |
| **Extreme capsule (L)** | 0.222 | 0.002 | 0.226 | 0.001 | -0.619 | 0.006 |
| **Extreme capsule (R)** | 0.224 | 0.001 | 0.227 | 0.001 | -0.462 | 0.032 |
| **Frontal aslant tract (L)** | 0.211 | 0.002 | 0.217 | 0.002 | -0.614 | 0.026 |
| **Frontal aslant tract (R)** | 0.213 | 0.002 | 0.219 | 0.002 | -0.662 | 0.016 |
| **Inferior longitudinal fasciculus (L)** | 0.228 | 0.002 | 0.231 | 0.001 | -0.575 | 0.013 |
| **Inferior longitudinal fasciculus (R)** | 0.226 | 0.001 | 0.229 | 0.001 | -0.592 | 0.008 |
| **Middle cerebellar peduncle** | 0.211 | 0.003 | 0.215 | 0.002 | -0.401 | 0.105 |
| **Middle longitudinal fasciculus (L)** | 0.220 | 0.002 | 0.224 | 0.001 | -0.688 | 0.006 |
| **Middle longitudinal fasciculus (R)** | 0.217 | 0.001 | 0.222 | 0.001 | -0.841 | 0.001 |
| **Optic radiation (L)** | 0.233 | 0.002 | 0.239 | 0.001 | -0.724 | 0.004 |
| **Optic radiation (R)** | 0.233 | 0.002 | 0.237 | 0.001 | -0.492 | 0.046 |
| **Superior longitudinal fasciculus I (L)** | 0.203 | 0.003 | 0.211 | 0.002 | -0.569 | 0.035 |
| **Superior longitudinal fasciculus I (R)** | 0.204 | 0.003 | 0.211 | 0.002 | -0.619 | 0.024 |
| **Superior longitudinal fasciculus II (L)** | 0.217 | 0.002 | 0.223 | 0.001 | -0.848 | 0.001 |
| **Superior longitudinal fasciculus II (R)** | 0.216 | 0.001 | 0.222 | 0.001 | -1.011 | 0.000 |
| **Superior longitudinal fasciculus III (L)** | 0.220 | 0.001 | 0.225 | 0.001 | -0.805 | 0.002 |
| **Superior longitudinal fasciculus III (R)** | 0.221 | 0.001 | 0.226 | 0.001 | -0.858 | 0.001 |
| **Uncinate fasciculus (L)** | 0.214 | 0.002 | 0.220 | 0.001 | -0.724 | 0.004 |
| **Uncinate fasciculus (R)** | 0.217 | 0.002 | 0.221 | 0.001 | -0.635 | 0.009 |

*CC = corpus callosum; d = Cohen’s d effect size (TD - RASopathies); FDR=false discovery rate; L = left; MTV = macromolecular tissue volume; R = right*

**Supplementary Table 2. Incorrectly classified RASopathies subjects following leave-one-out cross-validation, compared to the group averages of the correctly classified RASopathies subjects.**

| **Subject** | **Expressive language** | **Executive function** | **Receptive language** | **Inhibitory control and attention** | **Working memory** | **PreSubiculum (R)** | **Area 23d (R)** | **Area anterior 32 prime (R)** | **Superior frontal language area (L)** | **Area PH (R)** |
| --- | --- | --- | --- | --- | --- | --- | --- | --- | --- | --- |
| **Group average** | **90** | **88** | **98** | **81** | **94** | **0.623** | **0.662** | **0.637** | **0.586** | **0.608** |
| 1 | 90 | 94 | 102 | 83 | 107 | 0.646 | 0.650 | 0.663 | 0.618 | 0.610 |
| 3 | 87 | 80 | 113 | 88 | 83 | 0.672 | 0.663 | 0.645 | 0.594 | 0.620 |
| 2 | 118 | 80 | 97 | 100 | 100 | 0.622 | 0.645 | 0.629 | 0.629 | 0.600 |
| 5 | 106 | 111 | 106 | 101 | 77 | 0.603 | 0.649 | 0.664 | 0.617 | 0.599 |
| 4 | 83 | 82 | 102 | 84 | 90 | 0.633 | 0.652 | 0.630 | 0.641 | 0.611 |

These subjects were incorrectly classified as TD in the SVM following leave-one-out cross-validation. The group average refers to correctly classified RASopathies subjects.

**Supplementary Table 3. Incorrectly classified TD subjects following leave-one-out cross-validation, compared to the group averages of correctly classified TD subjects.**

| **Subject** | **Expressive language** | **Executive function** | **Receptive language** | **Inhibitory control and attention** | **Working memory** | **PreSubiculum (R)** | **Area 23d (R)** | **Area anterior 32 prime (R)** | **Superior frontal language area (L)** | **Area PH (R)** |
| --- | --- | --- | --- | --- | --- | --- | --- | --- | --- | --- |
| **Group average** | **114** | **112** | **119** | **99** | **94** | **0.602** | **0.669** | **0.642** | **0.599** | **0.620** |
| 1 | 100 | 133 | 100 | 92 | 89 | 0.606 | 0.701 | 0.660 | 0.608 | 0.621 |
| 2 | 89 | 103 | 140 | 90 | 121 | 0.613 | 0.658 | 0.652 | 0.587 | 0.637 |
| 3 | 90 | 94 | 122 | 83 | 93 | 0.615 | 0.693 | 0.663 | 0.597 | 0.624 |

TD subjects incorrectly classified in the RASopathies class in the SVM.

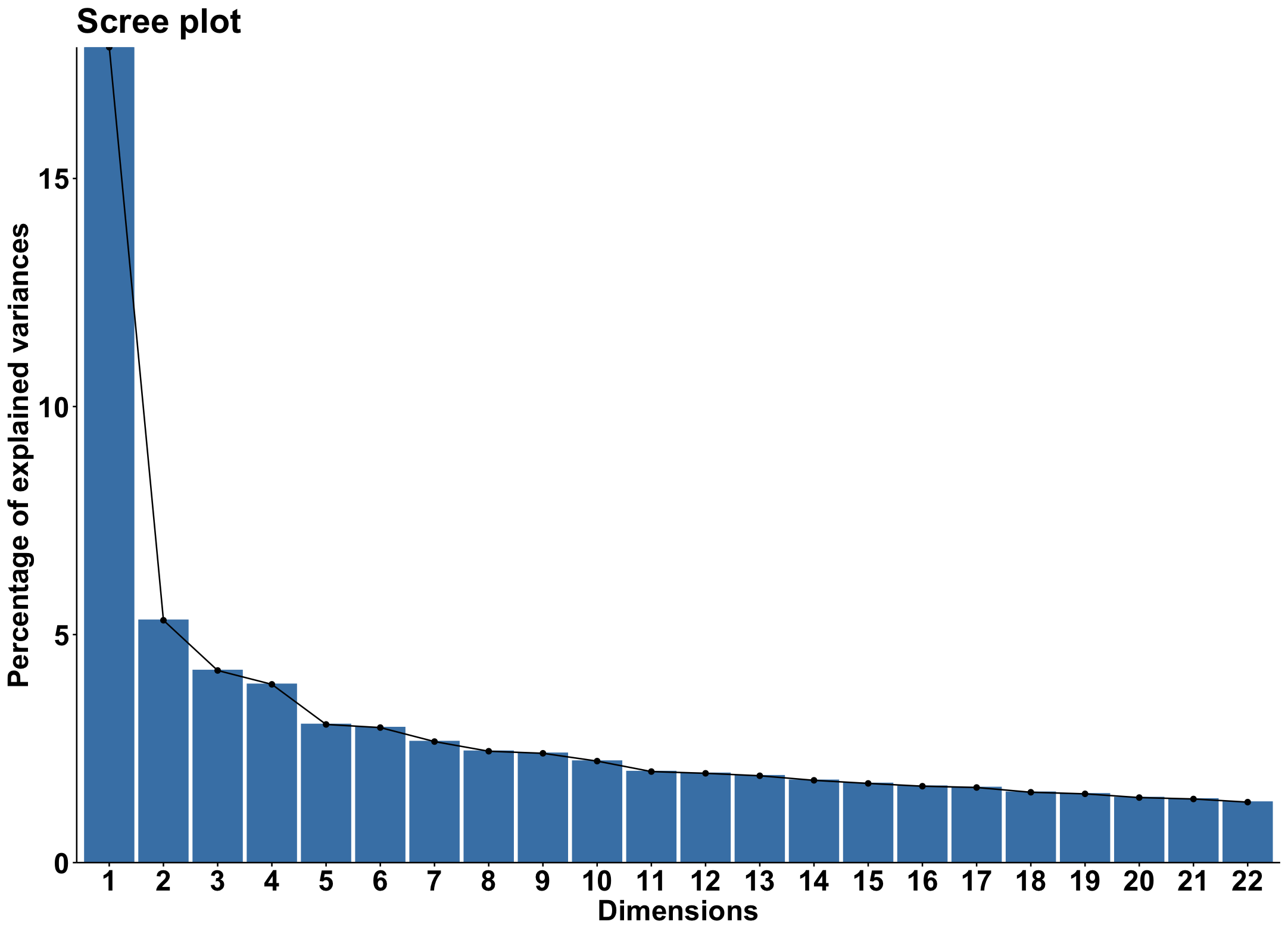

**Supplementary Figure 1. Scree plot of the percentage variance explained by each of the top 22 principal components which cumulatively explain 67% of the variance in the cortical R1 data.**

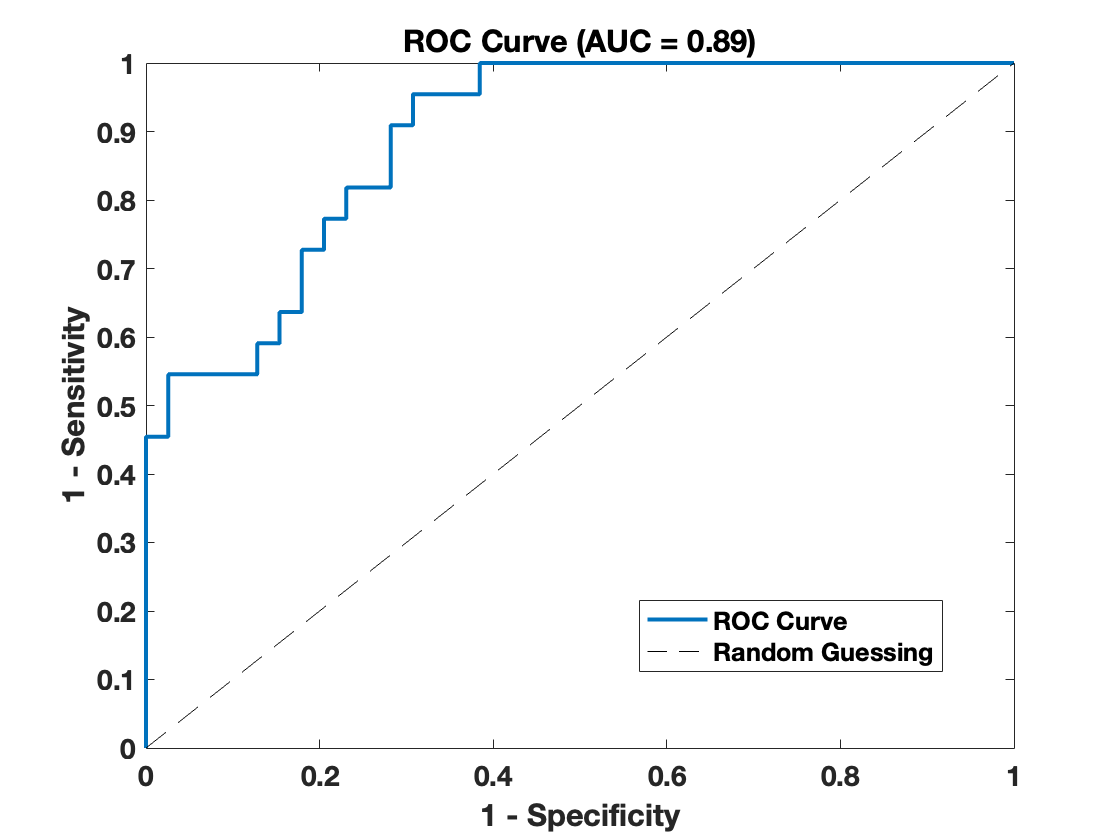

**Supplementary Figure 2. The ROC curve produced by the second SVM classifier. This SVM was generated based on cognitive features only, excluding brain-based features. The area under the curve (AUC) is 0.89.** The cognitive measures used to create the SVM included the following age-standardized tests: language (Oral Reading Recognition Test), inhibitory control and attention (Flanker Inhibitory Control and Attention Test), receptive language (Picture Vocabulary Test), executive function (Dimensional Change Card Sort Test), and working memory (List Sorting Working Memory Test) are derived from the NIH toolbox tests named in brackets. *L=left; R=right; ROC=receiver operating characteristic.*

**Supplementary References**

1.	Marshall WA, Tanner JM. Variations in the pattern of pubertal changes in boys. Arch Dis Child. 1970 Feb;45(239):13–23.

2.	Marshall WA, Tanner JM. Variations in pattern of pubertal changes in girls. Arch Dis Child. 1969 Jun;44(235):291–303.

3.	Wechsler D. Wechsler Abbreviated Scale of Intelligence [Internet]. 1999. Available from: https://psycnet.apa.org/fulltext/9999-15170-000.pdf

4.	Zelazo PD. The Dimensional Change Card Sort (DCCS): a method of assessing executive function in children. Nat Protoc. 2006;1(1):297–301.

5.	Zelazo PD, Anderson JE, Richler J, Wallner-Allen K, Beaumont JL, Weintraub S. II. NIH Toolbox Cognition Battery (CB): measuring executive function and attention. Monogr Soc Res Child Dev. 2013 Aug;78(4):16–33.

6.	Tulsky DS, Carlozzi NE, Chevalier N, Espy KA, Beaumont JL, Mungas D. V. NIH Toolbox Cognition Battery (CB): measuring working memory. Monogr Soc Res Child Dev. 2013 Aug;78(4):70–87.

7.	Tulsky DS, Carlozzi N, Chiaravalloti ND, Beaumont JL, Kisala PA, Mungas D, et al. NIH Toolbox Cognition Battery (NIHTB-CB): list sorting test to measure working memory. J Int Neuropsychol Soc. 2014 Jul;20(6):599–610.

8.	Gershon RC, Cook KF, Mungas D, Manly JJ, Slotkin J, Beaumont JL, et al. Language measures of the NIH Toolbox Cognition Battery. J Int Neuropsychol Soc. 2014 Jul;20(6):642–51.

9.	Gershon RC, Slotkin J, Manly JJ, Blitz DL, Beaumont JL, Schnipke D, et al. IV. NIH Toolbox Cognition Battery (CB): measuring language (vocabulary comprehension and reading decoding). Monogr Soc Res Child Dev. 2013 Aug;78(4):49–69.

10.	Carlozzi NE, Tulsky DS, Chiaravalloti ND, Beaumont JL, Weintraub S, Conway K, et al. NIH Toolbox Cognitive Battery (NIHTB-CB): the NIHTB Pattern Comparison Processing Speed Test. J Int Neuropsychol Soc. 2014 Jul;20(6):630–41.

11.	Dikmen SS, Bauer PJ, Weintraub S, Mungas D, Slotkin J, Beaumont JL, et al. Measuring episodic memory across the lifespan: NIH Toolbox Picture Sequence Memory Test. J Int Neuropsychol Soc. 2014 Jul;20(6):611–9.
